# Persistent high mortality rates for Diabetes Mellitus and Hypertension after excluding deaths associated with COVID-19 in Brazil, 2020-2022

**DOI:** 10.1101/2023.10.17.23297174

**Authors:** Rodrigo Moreira, Leonardo S Bastos, Luiz Max Carvalho, Laís Picinini Freitas, Antonio G Pacheco

## Abstract

**Background:** The outbreak of severe acute respiratory syndrome coronavirus 2 (SARS-CoV-2) posed a significant public health challenge globally, with Brazil being no exception. Excess mortality during this period reached alarming levels. Cardiovascular diseases (CVD), Systemic Hypertension (HTN), and Diabetes Mellitus (DM) were associated with increased mortality. However, the specific impact of DM and HTN on mortality during the pandemic remains poorly understood.

**Methods:** This study analyzed mortality data from Brazil’s mortality system, covering the period from 2015 to 2022. Data included all causes of death as listed on death certificates, categorized by International Classification of Diseases 10th edition (ICD-10) codes. Population data were obtained from the Brazilian Census. Mortality ratios (MRs) were calculated by comparing death rates in 2020, 2021, and 2022 to the average rates from 2015 to 2019. Adjusted MRs were calculated using Poisson models.

**Results:** Between 2015 and 2022, Brazil recorded a total of 11,423,288 deaths. Death rates remained relatively stable until 2019 but experienced a sharp increase in 2020 and 2021. In 2022, although a decrease was observed, it did not return to pre-pandemic levels. This trend persisted even when analyzing records mentioning DM, HTN, or CVD. Excluding death certificates mentioning COVID-19 codes, the trends still showed increases from 2020 through 2022, though less pronounced.

**Conclusion:** This study highlights the persistent high mortality rates for DM and HTN in Brazil during the years 2020-2022, even after excluding deaths associated with COVID-19. These findings emphasize the need for continued attention to managing and preventing DM and HTN as part of public health strategies, both during and beyond the COVID-19 pandemic. There are complex interactions between these conditions and the pandemic’s impact on mortality rates.

## Introduction

Severe acute respiratory syndrome coronavirus 2 (SARS-CoV-2) disease (COVID-19) had been a major public health emergency worldwide and in Brazil (1), with high burden, hitting the hardest in 2020 and 2021. Excess mortality during that period reached very high values in many countries. One study pointed to excess rates as high as 734.9 per 100,000 inhabitants in Bolivia (2). In Brazil, even though excess mortality was heterogeneous among states of residence, a rate of 186.9 per 100.000 was reported in that same study. Other studies reported excess deaths ranging from 10% to 40% in that same period in Brazil (3–5).

Cardiovascular diseases (CVD) and associated conditions such as Systemic Hypertension (HTN) and Diabetes Mellitus (DM) have been associated with severe COVID-19 and mortality (1, 6, 7), even though the role of HTN and DM as independent risk factors are not yet clear (8).

Excess mortality and increased rates of CVD mortality have been reported worldwide and in Brazil (9, 10), but detailed data for DM and HTN as contributing morbidities during the pandemic is scarce.

In this study we compared sex, age and state of residence adjusted mortality ratios (aMR) in Brazil in 2020-2022, compared to the preceding period of 2015-2019 and in subgroups of CVD, DM and HTN whenever these conditions were mentioned on death certificates. Comparisons were made with and without COVID-19 mentioned on the death certificates.

## Methods

Mortality data used in this study comes from the Brazilian mortality system (SIM - *Sistema de Informação sobre Mortalidade*) and is publicly available from DATASUS (Opendatasus - https://opendatasus.saude.gov.br/dataset/sim). Files for all-cause mortality, including multiple causes as assigned on death certificates, from january 2015 through december 2022, were downloaded and processed as described below. The 2022 database was deemed as preliminary data at the time of this analysis (accessed on 08/30/2023).

All data from death certificates except information that could identify individuals were available for analysis, including all causes of death (CoD) mentioned on death certificates and the underlying cause of death, which is a calculated variable, based on the information of immediate, contributing and concomitant causes leading to death. All causes are coded into International Classification of Diseases 10th. edition (ICD-10) codes and can thus be grouped according to what is being studied.

Population data were obtained from the Brazilian Census Bureau (IBGE – *Instituto Brasileiro de Geografia e Estatística*) through the SIDRA system (http://api.sidra.ibge.gov.br/). We obtained population projections per age group, sex and state of residence from january 2015 to december 2022.

For this study, we worked with ICD-10 codes of interest mentioned in any field from the death certificates. The following groups were created:

- COVID-19 – ICDs: B342, U071, U072

- Diabetes Mellitus (DM) – ICDs: E10 through E14

- High blood pressure (HBP) – ICDs: I10 through I15

- CVD: Cardiovascular diseases – ICDs: I00 through I99, except I46 (cardiac arrest)

Mortality ratios (MRs) were calculated as the death rates in 2020, 2021 and 2022 over the average death rates from 2015 through 2019. Values above one were considered excess when compared to the non-pandemic period. The numerator of the rates included all death certificates that mentioned any ICD-10 codes described above in any field of the death certificate. To measure the impact of COVID-19, a new rate, excluding death certificates that also mentioned COVID-19 in any field were excluded.

Adjusted MRs were calculated through Poisson models using the log of the population as an offset. All models were adjusted for state of residence, age group and sex. When controlling for state of residence, mixed-effect models with random intercepts were used.

Ethical statement: Our research uses publicly available Information and aggregated data without individual identification (DataSUS and IBGE data). Thus, an exemption from submission to the Institutional Review Board is provided by federal Brazilian resolution, CNS n.° 510, from 2016.

All analyses were performed with R 4.2.2 (11).

## Results

A total of 11,423,288 deaths were recorded in Brazil from 2015 to 2022. The rates were fairly stable over time until 2019, after which there was a steep rise in 2020 and 2021. In 2022 there was a decrease, but not to the same level as before (Figure 1). This pattern was also noticed when only records that mentioned DM, HTN or CVD were selected. When records that also mentioned COVID-19 codes were excluded, all trends still presented increases from 2020 through 2022, though much smoother (Figure 1). Overall characteristics of deaths in Brazil are depicted in Supplementary Table 1.

**Figure 1.**
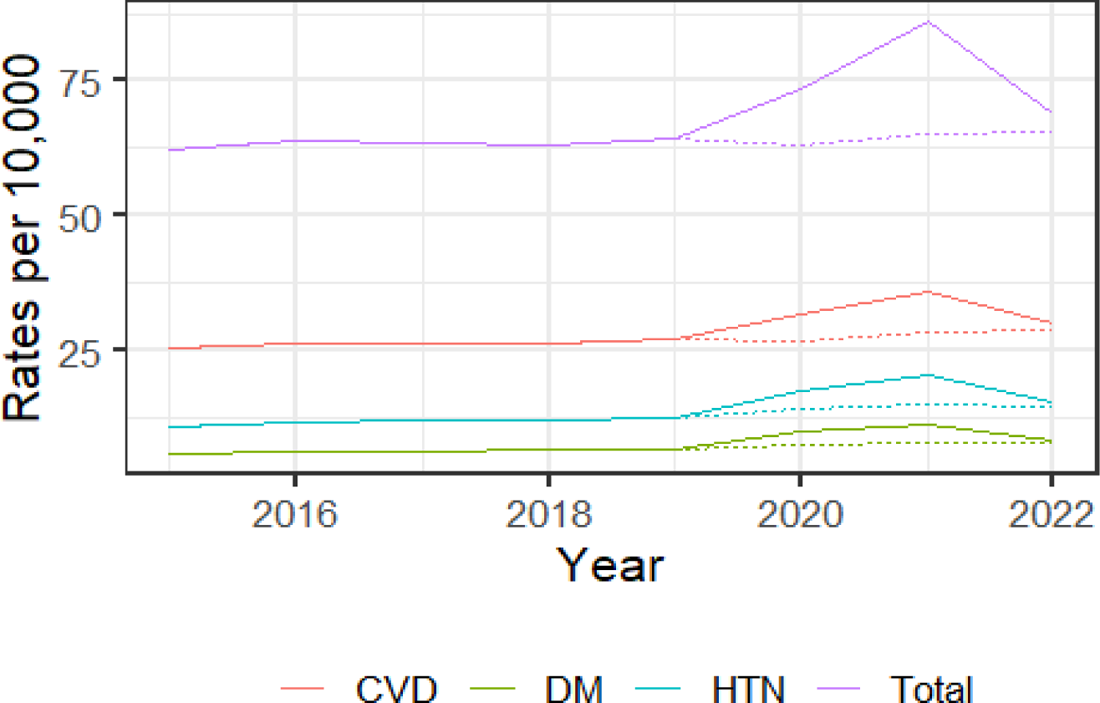
Mortality rates in Brazil with (solid lines) and without (dotted lines) COVID-19 mentioned in the death certificate - Overall, Cardiovascular Disease (CVD), Diabetes Mellitus (DM) and Hypertension (HTN) - 2015-2022)

Overall, adjusted mortality ratios were 9% and 24% higher for 2020 and 2021, respectively, compared to 2015-2019, whereas for 2022 it was 2% lower. For all three years, when COVID-19 was removed from the numerator, total aMRs fell back to values around 0.93. A similar configuration is noted for CVD mortality rates (Table 1).

**Table 1.** Adjusted Mortality Ratios (95% Confidence Intervals) comparisons among individuals who had Diabetes Mellitus (DM), Hypertension (HTN) or Cardiovascular Disease (CVD) mentioned in their death certificate with and without mention of COVID-19.

| Category | 2020 | 2021 | 2022 |
| --- | --- | --- | --- |
| Total | 1.09 (1.08-1.09) | 1.24 (1.24-1.25) | 0.98 (0.97-0.98) |
| Total, no COVID | 0.93 (0.93-0.93) | 0.94 (0.94-0.94) | 0.93 (0.93-0.93) |
| DM | 1.45 (1.44-1.45) | 1.61 (1.6-1.61) | 1.14 (1.13-1.14) |
| DM, no COVID | 1.12 (1.11-1.12) | 1.15 (1.14-1.15) | 1.07 (1.06-1.07) |
| HTN | 1.37 (1.36-1.37) | 1.54 (1.54-1.55) | 1.14 (1.14-1.15) |
| HTN, no COVID | 1.11 (1.11-1.12) | 1.15 (1.15-1.15) | 1.09 (1.08-1.09) |
| CVD | 1.1 (1.1-1.1) | 1.23 (1.23-1.23) | 0.99 (0.99-1) |
| CVD, no COVID | 0.94 (0.94-0.94) | 0.97 (0.97-0.98) | 0.95 (0.95-0.95) |

When we look at mortality rates for DM and HTN, aMRs are higher than those for overall and CVD (ranging from 1.14 in 2022 to 1.61 in 2021 for DM and 1.14 in 2022 to 1.54 in 2021), but they did not fall below 1 when COVID-19 records are excluded, reaching a 15% increase for both conditions in 2021 (Table 1).

Those figures were not homogeneous when we look within age subgroups either. As shown in Figure 2, significant increased ratios were noted from 30 years and older for all groups in 2020 and 2021 but not in 2022, when looking at all causes of death (Panel A). In 2021 middle-aged adults were particularly impacted with a 48% increase in overall mortality in the 40–49-year-old group. All ratios return to less than 1 when COVID-19 is removed. A similar pattern was noted for CVD (Panel D). For DM (Panel B) and HTN (Panel C), not only does the increase in the ratios begin in younger age groups (0-19 and 20-29, respectively), but also they did not return to baseline values when death certificates that mentioned COVID-19 were removed from the analysis. Similar patterns are noted for sex (Supplementary Figure 1).

**Figure 2.**
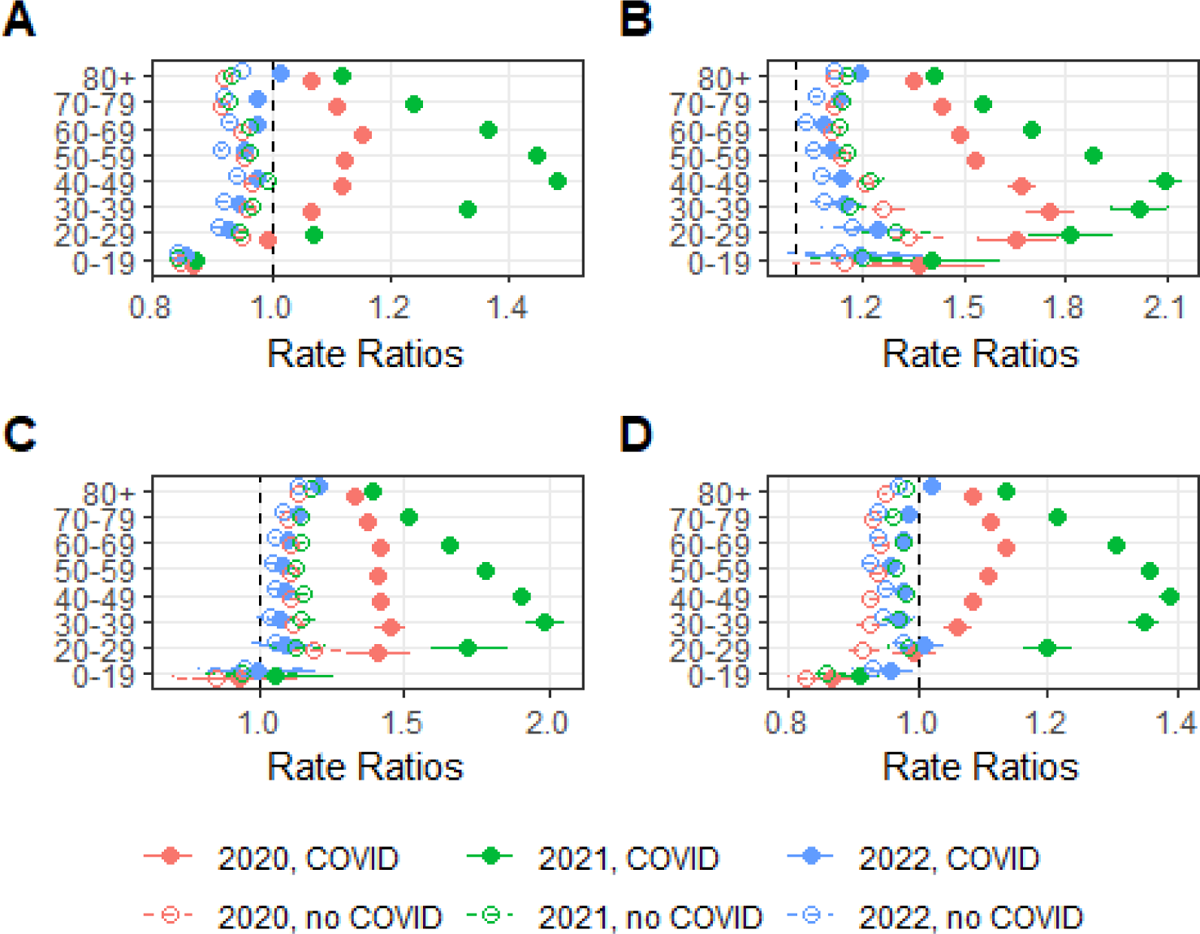
Adjusted Mortality Ratios in Brazil with and without COVID-19 mentioned in the death certificate per age group – A - Overall, B – DM, C – HTN, D – CVD; baseline - 2015-2019)

Across states, figures were heterogeneous. In 2020 overall mortality rates increased for most states (Supplementary Figure 2), ranging from −2% in Rio Grande do Sul to 30% increase in Amazonas, whereas in 2021 it ranged from 12% (Alagoas and Sergipe) up to 49% in Rondônia. Of note, Amazonas, one of the states with the deadliest outcomes during the pandemic experienced a 47% increase rate in 2021 on top of that 30% in 2020. All ratios also returned to baseline values upon the removal of COVID-19. In 2022 some states did have significantly increased ratios, but all of them returned to values below 1 when COVID-19 was removed. Similar patterns were observed for CVD (Supplementary Figure 5). Regarding DM and HTN (Supplementary Figures 3 and 4) it followed the same trends as the overall mortality ratios, once again with AM displaying higher aMR among all the states.

## Discussion

In this study we showed increased mortality rates during the COVID-19 pandemic in Brazil among deceased individuals that had CVD, HTN and DM mentioned in their death certificates. Even though those are expected results, rates did not return to baseline values after removing cases with concomitant COVID-19 for DM and HTN, as happened with overall (total) and CVD subgroup. Our results are in line with the literature, in terms of the overall and CVD mortality ratios.

In Brazil, overall excess mortality ranged from 10% to 40%, depending on the study and period studied (3–5), which is close to our mortality ratios in 2020 (9%) and 2021 (24%). If we did look at excess mortality, our data would show figures even closer to those (14% and 32.4%, respectively – data not shown). CVD mortality rates were about the same level as overall rates and are also in line with the literature (9).

It is worth highlighting that Brazil faced many challenges in dealing with COVID-19, at some point having the highest number of cases and deaths in Latin America (12).. Among the regions most impacted was the state of Amazonas which experienced the highest mortality rates in the country during the pandemic. Known disparities in healthcare access, overwhelming of the healthcare system and quality of care during this period, leading to delayed or inadequate treatment for COVID-19 might have contributed to such results.

We showed higher mortality rates from CVD in the presence of COVID-19 in this study. Viral infection can initiate myocardial injury and provoke inflammatory hyperactivity. Among the most prevalent cardiovascular complications observed in COVID-19 are myocardial infarction, myocarditis accompanied by reduced systolic function of the left ventricle, arrhythmias and thromboembolic complications (13).

As expected, death rates for those with HTN and DM as comorbidities also significantly increased in 2020 and 2021, but contrary to overall deaths and CVD, they did not return to baseline values after death certificates that mentioned COVID-19 were removed. Two different mechanisms may be playing a role on this pattern. The first one is the possibility of underreporting of COVID-19 among deceased individuals with those conditions in Brazil, during that period. Even though this mechanism is probably present in this case, and has been reported in Brazil (14), we don’t think this is the only factor playing a role here. First because overall and CVD death rates returned to baseline, and even to values lower than those in 2015-2019, since it is expected that mortality by causes not related to COVID-19 would decrease (14), and, This is especially true, because we removed all death certificates that mentioned COVID-19 and not only those with COVID-19 as the underlying cause of death, which would account for cases where misclassification occurred.

The other mechanism would be that these conditions in fact had their prevalence increased during the pandemic, and in fact contributed to increased death rates during this period, and even in 2022, as shown. The relationship of DM and HTN with life-style changes during the pandemic and even after it (e.g. decreased physical activity, poor glycemic control and adherence to therapy) may have had an impact on aMR in the population without COVID-19 (15). Also, some studies pointed to newly diagnosed HTN and DM after acute COVID-19 diagnosis, especially in severe cases (16, 17).

One possibility both for risk factors and increased prevalence would be the fact that most individuals with those conditions are of older age and would be driving this trend, as their death rates are higher than younger individuals [8]. Our results were adjusted for age, sex and state of residence, which should limit the impact of those distributions across groups. Moreover, we showed that rates in all age groups above 20-29 years have the same behavior, with a predominance among young adults (Figure 2).

This study has several limitations. First, it is based on data from death certificates, which are known to suffer from misclassification, despite going through thorough revision before information is entered into the database (18).. This problem results in both underreporting of conditions (especially for garbage ICD-10 codes are reported as underlying cause of death) or reporting wrong diagnoses. The approach used in this study to search for all causes mentioned tends to lower this problem and has been used before to study causes of death in Brazil (19)..

Another limitation is the number of variables used to adjust the rates. Even though race/ethnicity, education and other characteristics are also available on death certificates, there are difficulties in respect to missing values and population projection for those subgroups. Since in our case we wanted to control for major factors that would impact on HTN and DM rates, we believe that sex, age and state of residence would cover most heterogeneities involved in those calculations.

Strong points of our study include the use of a large, representative database in Brazil, that comprises all deaths reported to the Ministry of Health and the possibility to study multiple causes of death instead of the underlying causes, as is usually done in mortality studies.

In conclusion, our study showed persistent higher mortality in individuals with a diagnosis of HTN and DM at the time of death in Brazil during the COVID-19 pandemic, even after removing those deaths related to COVID-19. This finding should point to improving diagnosis of COVID-19 and correctly reporting it on the death certificate, and also increase surveillance for both HTN and DM in patients who recently had a COVID-19 diagnosis to better control those conditions among them.

## Data Availability

Data will be available upon request

http://api.sidra.ibge.gov.br/

## Supporting information

**Supplementary Figure 1.**
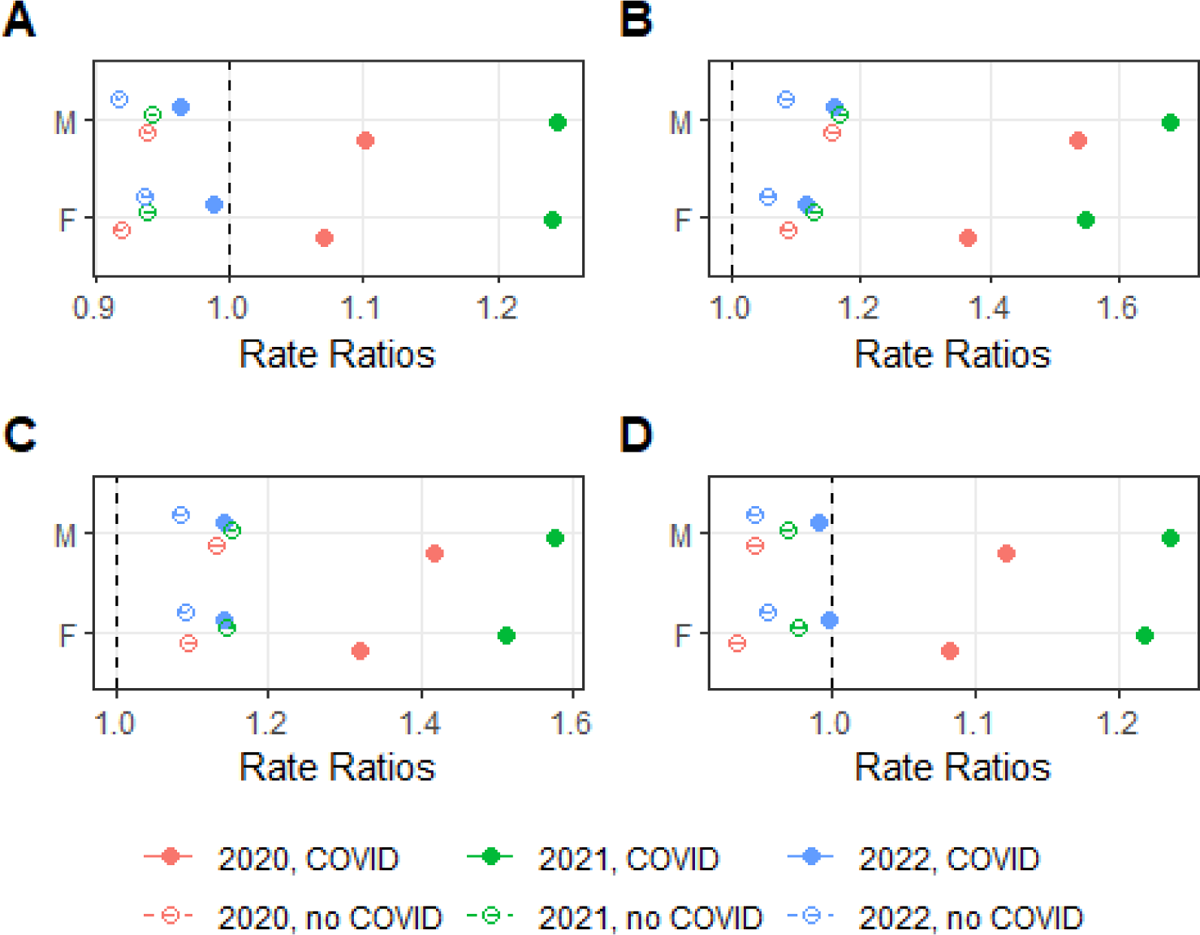
Adjusted Mortality Ratios in Brazil with and without COVID-19 mentioned in the death certificate per age group – A - Overall, B – DM, C – HTN, D – CVD; baseline - 2015-2019)

**Supplementary Figures 2 - 5.**
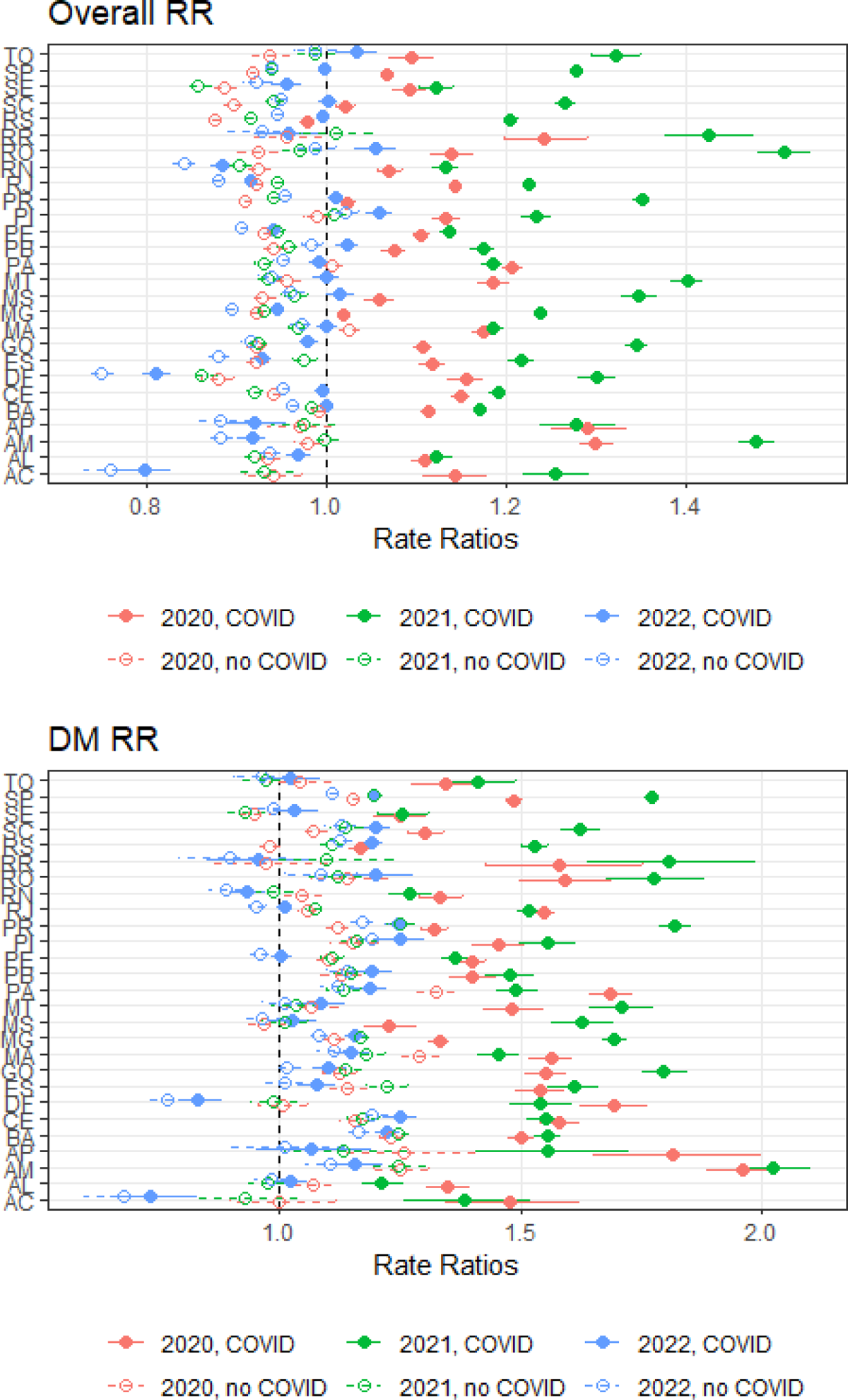
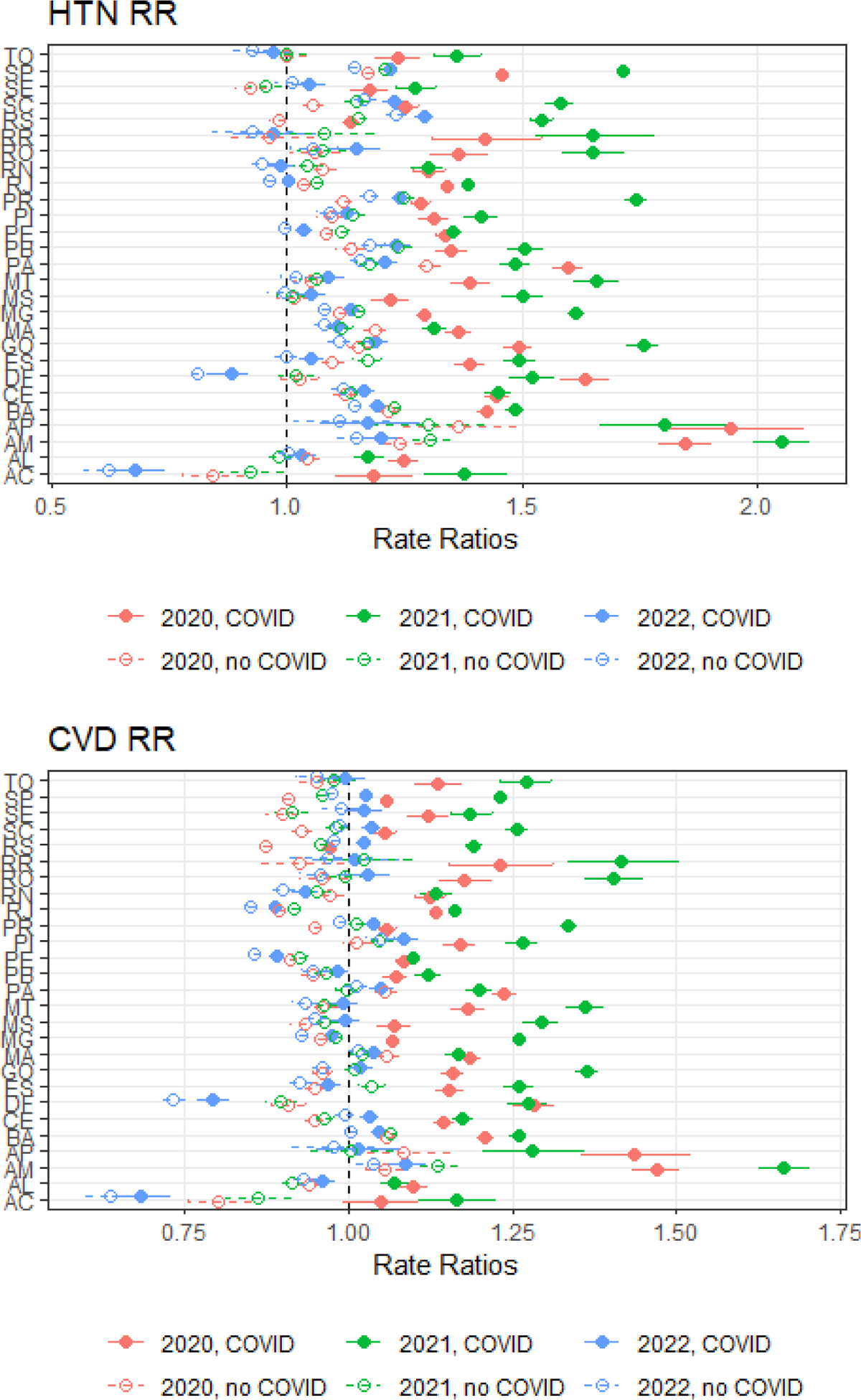
State of Residence

---

**Supplementary Table 1.** Characteristics of mortality data in Brazil, 2015-2022.

| **Variables** | **2015** | **2016** | **2017** | **2018** | **2019** | **2020** | **2021** | **2022** | **Test** | **P-value** |
| --- | --- | --- | --- | --- | --- | --- | --- | --- | --- | --- |
| *Total* | 1264<br>175 | 1309<br>774 | 1312<br>663 | 1316<br>719 | 1349<br>801 | 1556<br>824 | 1832<br>649 | 1480<br>683 |  |  |
| **Sex** |  |  |  |  |  |  |  |  | Chisq.<br>(7 df)<br>= 1315.<br>92 | <<br>0.0<br>01 |
| *F* | 5543<br>83<br>(43.9<br>) | 5723<br>59<br>(43.7<br>) | 5775<br>73<br>(44) | 5824<br>57<br>(44.3<br>) | 6037<br>25<br>(44.7<br>) | 6820<br>27<br>(43.8<br>) | 8166<br>16<br>(44.6<br>) | 6716<br>39<br>(45.4<br>) |  |  |
| *M* | 7091<br>17<br>(56.1<br>) | 7368<br>42<br>(56.3<br>) | 7344<br>69<br>(56) | 7336<br>16<br>(55.7<br>) | 7455<br>19<br>(55.3<br>) | 8741<br>67<br>(56.2<br>) | 1015<br>350<br>(55.4<br>) | 8084<br>31<br>(54.6<br>) |  |  |
| **Age group** |  |  |  |  |  |  |  |  | Chisq.<br>(49<br>df) = 4913<br>9.97 | <<br>0.0<br>01 |
| *0-19* | 7261<br>1<br>(5.8) | 7252<br>4<br>(5.6) | 7151<br>7<br>(5.5) | 6827<br>0<br>(5.2) | 6520<br>6<br>(4.8) | 5912<br>9<br>(3.8) | 5902<br>6<br>(3.2) | 5773<br>6<br>(3.9) |  |  |
| *20-29* | 5433<br>6<br>(4.3) | 5564<br>3<br>(4.3) | 5575<br>5<br>(4.3) | 5195<br>2 (4) | 4886<br>2<br>(3.6) | 5295<br>3<br>(3.4) | 5683<br>6<br>(3.1) | 4889<br>0<br>(3.3) |  |  |
| *30-39* | 6419<br>0<br>(5.1) | 6486<br>4 (5) | 6377<br>6<br>(4.9) | 6115<br>1<br>(4.7) | 5998<br>4<br>(4.5) | 6816<br>4<br>(4.4) | 8520<br>2<br>(4.7) | 6046<br>8<br>(4.1) |  |  |
| *40-49* | 9099<br>3<br>(7.2) | 9265<br>0<br>(7.1) | 8917<br>7<br>(6.8) | 8882<br>6<br>(6.8) | 8895<br>7<br>(6.6) | 1064<br>88<br>(6.9) | 1439<br>75<br>(7.9) | 9653<br>6<br>(6.5) |  |  |
| *50-59* | 1523<br>31 | 1577<br>97 | 1532<br>93 | 1544<br>01 | 1554<br>34 | 1841<br>02 | 2406<br>90 | 1603<br>19 |  |  |
|  | (12.1<br>) | (12.1<br>) | (11.7<br>) | (11.8<br>) | (11.5<br>) | (11.8<br>) | (13.2<br>) | (10.8<br>) |  |  |
| 60-69 | 2096<br>20<br>(16.6<br>) | 2217<br>52<br>(17) | 2232<br>84<br>(17) | 2291<br>56<br>(17.4<br>) | 2368<br>22<br>(17.6<br>) | 2861<br>90<br>(18.4<br>) | 3501<br>31<br>(19.1<br>) | 2582<br>28<br>(17.5<br>) |  |  |
| 70-79 | 2561<br>80<br>(20.3<br>) | 2652<br>20<br>(20.3<br>) | 2670<br>12<br>(20.4<br>) | 2719<br>19<br>(20.7<br>) | 2820<br>40<br>(20.9<br>) | 3347<br>33<br>(21.5<br>) | 3900<br>64<br>(21.3<br>) | 3204<br>58<br>(21.7<br>) |  |  |
| 80+ | 3606<br>34<br>(28.6<br>) | 3761<br>45<br>(28.8<br>) | 3858<br>82<br>(29.5<br>) | 3882<br>71<br>(29.5<br>) | 4101<br>38<br>(30.4<br>) | 4626<br>78<br>(29.8<br>) | 5043<br>48<br>(27.6<br>) | 4759<br>25<br>(32.2<br>) |  |  |
| **DM** |  |  |  |  |  |  |  |  | Chisq.<br>(7 df)<br>=<br>2723<br>7.77 | <<br>0.0<br>01 |
| No | 1142<br>083<br>(90.3<br>) | 1181<br>838<br>(90.2<br>) | 1179<br>632<br>(89.9<br>) | 1178<br>859<br>(89.5<br>) | 1206<br>536<br>(89.4<br>) | 1343<br>735<br>(86.3<br>) | 1587<br>388<br>(86.6<br>) | 1301<br>339<br>(87.9<br>) |  |  |
| Yes | 1220<br>92<br>(9.7) | 1279<br>36<br>(9.8) | 1330<br>31<br>(10.1<br>) | 1378<br>60<br>(10.5<br>) | 1432<br>65<br>(10.6<br>) | 2130<br>89<br>(13.7<br>) | 2452<br>61<br>(13.4<br>) | 1793<br>44<br>(12.1<br>) |  |  |
| **HBP** |  |  |  |  |  |  |  |  | Chisq.<br>(7 df)<br>=<br>4144<br>1.79 | <<br>0.0<br>01 |
| No | 1038<br>326<br>(82.1<br>) | 1071<br>858<br>(81.8<br>) | 1063<br>293<br>(81) | 1061<br>823<br>(80.6<br>) | 1085<br>979<br>(80.5<br>) | 1182<br>248<br>(75.9<br>) | 1394<br>843<br>(76.1<br>) | 1144<br>904<br>(77.3<br>) |  |  |
| Yes | 2258<br>49<br>(17.9<br>) | 2379<br>16<br>(18.2<br>) | 2493<br>70<br>(19) | 2548<br>96<br>(19.4<br>) | 2638<br>22<br>(19.5<br>) | 3745<br>76<br>(24.1<br>) | 4378<br>06<br>(23.9<br>) | 3357<br>79<br>(22.7<br>) |  |  |
| **CVD** |  |  |  |  |  |  |  |  | Chisq.<br>(7 df)<br>= | <<br>0.0<br>01 |
|  |  |  |  |  |  |  |  |  | 2234.<br>39 |  |
| *No* | 7401<br>73<br>(58.5<br>) | 7647<br>16<br>(58.4<br>) | 7648<br>23<br>(58.3<br>) | 7623<br>19<br>(57.9<br>) | 7810<br>90<br>(57.9<br>) | 8875<br>65<br>(57) | 1060<br>667<br>(57.9<br>) | 8345<br>80<br>(56.4<br>) |  |  |
| *Yes* | 5240<br>02<br>(41.5<br>) | 5450<br>58<br>(41.6<br>) | 5478<br>40<br>(41.7<br>) | 5544<br>00<br>(42.1<br>) | 5687<br>11<br>(42.1<br>) | 6692<br>59<br>(43) | 7719<br>82<br>(42.1<br>) | 6461<br>03<br>(43.6<br>) |  |  |
| **State**<br>**of**<br>**resid**<br>**ence** |  |  |  |  |  |  |  |  | Chisq.<br>(182<br>df) =<br>1551<br>0.2 | <<br>0.0<br>01 |
| *AC* | 3517<br>(0.3) | 3763<br>(0.3) | 3832<br>(0.3) | 4094<br>(0.3) | 4098<br>(0.3) | 4860<br>(0.3) | 5496<br>(0.3) | 3607<br>(0.2) |  |  |
| *AL* | 1975<br>6<br>(1.6) | 2076<br>9<br>(1.6) | 2067<br>3<br>(1.6) | 1941<br>1<br>(1.5) | 2028<br>7<br>(1.5) | 2414<br>8<br>(1.6) | 2509<br>0<br>(1.4) | 2223<br>8<br>(1.5) |  |  |
| *AM* | 1667<br>5<br>(1.3) | 1679<br>9<br>(1.3) | 1728<br>1<br>(1.3) | 1771<br>0<br>(1.3) | 1832<br>7<br>(1.4) | 2476<br>5<br>(1.6) | 2908<br>0<br>(1.6) | 1869<br>6<br>(1.3) |  |  |
| *AP* | 2946<br>(0.2) | 2995<br>(0.2) | 3158<br>(0.2) | 3345<br>(0.3) | 3524<br>(0.3) | 4617<br>(0.3) | 4750<br>(0.3) | 3565<br>(0.2) |  |  |
| *BA* | 8708<br>3<br>(6.9) | 8809<br>4<br>(6.7) | 9091<br>5<br>(6.9) | 9013<br>4<br>(6.8) | 9336<br>5<br>(6.9) | 1071<br>94<br>(6.9) | 1153<br>92<br>(6.3) | 1006<br>29<br>(6.8) |  |  |
| *CE* | 5525<br>8<br>(4.4) | 5427<br>6<br>(4.1) | 5926<br>3<br>(4.5) | 5702<br>8<br>(4.3) | 5658<br>0<br>(4.2) | 6951<br>2<br>(4.5) | 7368<br>3 (4) | 6294<br>2<br>(4.3) |  |  |
| *DF* | 1197<br>5<br>(0.9) | 1205<br>0<br>(0.9) | 1251<br>4 (1) | 1215<br>7<br>(0.9) | 1280<br>4<br>(0.9) | 1621<br>8 (1) | 1907<br>9 (1) | 1243<br>3<br>(0.8) |  |  |
| *ES* | 2233<br>2<br>(1.8) | 2286<br>8<br>(1.7) | 2411<br>2<br>(1.8) | 2350<br>0<br>(1.8) | 2443<br>1<br>(1.8) | 2911<br>1<br>(1.9) | 3280<br>1<br>(1.8) | 2581<br>8<br>(1.7) |  |  |
| *GO* | 3885<br>4<br>(3.1) | 3807<br>4<br>(2.9) | 3997<br>3 (3) | 3950<br>7 (3) | 4102<br>5 (3) | 4835<br>8<br>(3.1) | 6071<br>2<br>(3.3) | 4573<br>1<br>(3.1) |  |  |
| *MA* | 3366<br>6<br>(2.7) | 3436<br>2<br>(2.6) | 3527<br>5<br>(2.7) | 3452<br>5<br>(2.6) | 3512<br>8<br>(2.6) | 4327<br>1<br>(2.8) | 4465<br>4<br>(2.4) | 3851<br>0<br>(2.6) |  |  |
| *MG* | 1312<br>74<br>(10.4<br>) | 1352<br>57<br>(10.3<br>) | 1381<br>18<br>(10.5<br>) | 1356<br>19<br>(10.3<br>) | 1410<br>22<br>(10.4<br>) | 1521<br>28<br>(9.8) | 1900<br>85<br>(10.4<br>) | 1494<br>49<br>(10.1<br>) |  |  |
| *MS* | 1545<br>7<br>(1.2) | 1674<br>9<br>(1.3) | 1595<br>4<br>(1.2) | 1660<br>0<br>(1.3) | 1681<br>5<br>(1.2) | 1905<br>1<br>(1.2) | 2504<br>9<br>(1.4) | 1947<br>6<br>(1.3) |  |  |
| *MT* | 1709<br>5<br>(1.4) | 1753<br>5<br>(1.3) | 1770<br>9<br>(1.3) | 1820<br>5<br>(1.4) | 1834<br>1<br>(1.4) | 2339<br>7<br>(1.5) | 2862<br>3<br>(1.6) | 2113<br>6<br>(1.4) |  |  |
| *PA* | 3736<br>5 (3) | 3855<br>7<br>(2.9) | 3998<br>0 (3) | 4051<br>3<br>(3.1) | 4059<br>9 (3) | 5164<br>3<br>(3.3) | 5216<br>6<br>(2.8) | 4501<br>7 (3) |  |  |
| *PB* | 2642<br>2<br>(2.1) | 2804<br>1<br>(2.1) | 2697<br>5<br>(2.1) | 2664<br>4 (2) | 2737<br>8 (2) | 3110<br>7 (2) | 3467<br>8<br>(1.9) | 3088<br>8<br>(2.1) |  |  |
| *PE* | 6255<br>6<br>(4.9) | 6692<br>8<br>(5.1) | 6436<br>4<br>(4.9) | 6201<br>1<br>(4.7) | 6429<br>5<br>(4.8) | 7657<br>4<br>(4.9) | 8071<br>7<br>(4.4) | 6881<br>8<br>(4.6) |  |  |
| *PI* | 1936<br>6<br>(1.5) | 1918<br>7<br>(1.5) | 1985<br>0<br>(1.5) | 1998<br>3<br>(1.5) | 2052<br>8<br>(1.5) | 2364<br>6<br>(1.5) | 2621<br>2<br>(1.4) | 2290<br>6<br>(1.5) |  |  |
| *PR* | 7083<br>9<br>(5.6) | 7474<br>0<br>(5.7) | 7163<br>3<br>(5.5) | 7384<br>8<br>(5.6) | 7456<br>6<br>(5.5) | 8257<br>3<br>(5.3) | 1126<br>06<br>(6.1) | 8693<br>0<br>(5.9) |  |  |
| *RJ* | 1327<br>14<br>(10.5<br>) | 1410<br>89<br>(10.8<br>) | 1367<br>09<br>(10.4<br>) | 1407<br>06<br>(10.7<br>) | 1446<br>00<br>(10.7<br>) | 1721<br>85<br>(11.1<br>) | 1892<br>32<br>(10.3<br>) | 1453<br>37<br>(9.8) |  |  |
| *RN* | 2015<br>3<br>(1.6) | 2192<br>2<br>(1.7) | 2140<br>9<br>(1.6) | 2120<br>9<br>(1.6) | 2176<br>7<br>(1.6) | 2467<br>4<br>(1.6) | 2677<br>1<br>(1.5) | 2145<br>1<br>(1.4) |  |  |
| *RO* | 7948<br>(0.6) | 8344<br>(0.6) | 8219<br>(0.6) | 8165<br>(0.6) | 8338<br>(0.6) | 1027<br>8<br>(0.7) | 1404<br>2<br>(0.8) | 1013<br>0<br>(0.7) |  |  |
| *RR* | 2091<br>(0.2) | 2157<br>(0.2) | 2461<br>(0.2) | 2787<br>(0.2) | 2779<br>(0.2) | 3580<br>(0.2) | 4306<br>(0.2) | 3020<br>(0.2) |  |  |
| *RS* | 8234<br>9<br>(6.5) | 8758<br>3<br>(6.7) | 8624<br>1<br>(6.6) | 8861<br>8<br>(6.7) | 8923<br>8<br>(6.6) | 9279<br>1 (6) | 1177<br>22<br>(6.4) | 1001<br>42<br>(6.8) |  |  |
| *SC* | 3798<br>4 (3) | 4027<br>0<br>(3.1) | 3991<br>9 (3) | 4126<br>8<br>(3.1) | 4228<br>2<br>(3.1) | 4644<br>4 (3) | 5989<br>8<br>(3.3) | 4935<br>0<br>(3.3) |  |  |
| *SE* | 1345<br>3<br>(1.1) | 1351<br>6 (1) | 1332<br>1 (1) | 1302<br>4 (1) | 1347<br>3 (1) | 1579<br>3 (1) | 1667<br>5<br>(0.9) | 1460<br>5 (1) |  |  |
| *SP* | 2876<br>45<br>(22.8<br>) | 2963<br>59<br>(22.6<br>) | 2947<br>53<br>(22.5<br>) | 2983<br>13<br>(22.7<br>) | 3061<br>90<br>(22.7<br>) | 3496<br>35<br>(22.5<br>) | 4316<br>16<br>(23.6<br>) | 3485<br>91<br>(23.5<br>) |  |  |
| *TO* | 7402<br>(0.6) | 7490<br>(0.6) | 8052<br>(0.6) | 7795<br>(0.6) | 8021<br>(0.6) | 9271<br>(0.6) | 1151<br>4<br>(0.6) | 9268<br>(0.6) |  |  |

## References

1. Niquini RP, Lana RM, Pacheco AG, Cruz OG, Coelho FC, Carvalho LM, et al. Description and comparison of demographic characteristics and comorbidities in SARI from COVID-19, SARI from influenza, and the Brazilian general population. Cad Saude Publica. 2020;36(7):e00149420.

2. Collaborators C-EM. Estimating excess mortality due to the COVID-19 pandemic: a systematic analysis of COVID-19-related mortality, 2020-21. Lancet. 2022;399(10334):1513-36.

3. Nucci LB, Enes CC, Ferraz FR, da Silva IV, Rinaldi AEM, Conde WL. Excess mortality associated with COVID-19 in Brazil: 2020-2021. J Public Health (Oxf). 2023;45(1):e7–e9.

4. Silva GAE, Jardim BC, Santos C. Excess mortality in Brazil in times of Covid-19. Cien Saude Colet. 2020;25(9):3345–54.

5. Teixeira RA, Vasconcelos AMN, Torens A, Franca EB, Ishitani L, Bierrenbach AL, et al. Excess Mortality due to natural causes among whites and blacks during the COVID-19 pandemic in Brazil. Rev Soc Bras Med Trop. 2022;55(suppl 1):e0283.

6. Pavey H, Kulkarni S, Wood A, Ben-Shlomo Y, Sever P, McEniery C, et al. Primary hypertension, anti-hypertensive medications and the risk of severe COVID-19 in UK Biobank. PLoS One. 2022;17(11):e0276781.

7. Gupta A, Nayan N, Nair R, Kumar K, Joshi A, Sharma S, et al. Diabetes Mellitus and Hypertension Increase Risk of Death in Novel Corona Virus Patients Irrespective of Age: a Prospective Observational Study of Co-morbidities and COVID-19 from India. SN Compr Clin Med. 2021;3(4):937–44.

8. Yamazaki O, Shibata S. Severe COVID-19 and preexisting hypertension: a matter of age? Hypertens Res. 2022;45(9):1523–5.

9. Brant LCC, Nascimento BR, Teixeira RA, Lopes M, Malta DC, Oliveira GMM, et al. Excess of cardiovascular deaths during the COVID-19 pandemic in Brazilian capital cities. Heart. 2020;106(24):1898–905.

10. Palacio-Mejia LS, Hernandez-Avila JE, Hernandez-Avila M, Dyer-Leal D, Barranco A, Quezada-Sanchez AD, et al. Leading causes of excess mortality in Mexico during the COVID-19 pandemic 2020-2021: A death certificates study in a middle-income country. Lancet Reg Health Am. 2022;13:100303.

11. R Development Core Team. R: A language and environment for statistical computing. 4.2.2 ed. Vienna, Austria: R Foundation for Statistical Computing; 2023.

12. The L. COVID-19 in Brazil: “So what?”. Lancet. 2020;395(10235):1461.

13. Azevedo RB, Botelho BG, Hollanda JVG, Ferreira LVL, Junqueira de Andrade LZ, Oei S, et al. Covid-19 and the cardiovascular system: a comprehensive review. J Hum Hypertens. 2021;35(1):4–11.

14. Santos AMD, Souza BF, Carvalho CA, Campos MAG, Oliveira B, Diniz EM, et al. Excess deaths from all causes and by COVID-19 in Brazil in 2020. Rev Saude Publica. 2021;55:71.

15. Karagiannidis AG, Theodorakopoulou MP, Ferro CJ, Ortiz A, Soler MJ, Halimi JM, et al. Impact of public restrictive measures on hypertension during the COVID-19 pandemic: existing evidence and long-term implications. Clin Kidney J. 2023;16(4):619–34.

16. Zuin M, Rigatelli G, Bilato C, Pasquetto G, Mazza A. Risk of Incident New-Onset Arterial Hypertension After COVID-19 Recovery: A Systematic Review and Meta-analysis. High Blood Press Cardiovasc Prev. 2023;30(3):227–33.

17. Chourasia P, Goyal L, Kansal D, Roy S, Singh R, Mahata I, et al. Risk of New-Onset Diabetes Mellitus as a Post-COVID-19 Condition and Possible Mechanisms: A Scoping Review. J Clin Med. 2023;12(3).

18. Fajardo S, Aerts DR, Bassanesi SL. [Accuracy of the Mortality Information System team in the specification of underlying cause of death in a State capital in southern Brazil]. Cad Saude Publica. 2009;25(10):2218–28.

19. Paula AA, Schechter M, Tuboi SH, Faulhaber JC, Luz PM, Veloso VG, et al. Continuous increase of cardiovascular diseases, diabetes, and non-HIV related cancers as causes of death in HIV-infected individuals in Brazil: an analysis of nationwide data. PLoS One. 2014;9(4):e94636.

